# Heterogeneity in vaccinal immunity to SARS-CoV-2 can be addressed by a personalized booster strategy

**DOI:** 10.1101/2022.08.30.22279397

**Authors:** Madison Stoddard, Lin Yuan, Sharanya Sarkar, Shruthi Mangalaganesh, Ryan P. Nolan, Dean Bottino, Greg Hather, Natasha S. Hochberg, Laura F. White, Arijit Chakravarty

## Abstract

The ongoing COVID-19 pandemic has placed an unprecedented burden on global health. Crucial for managing this burden, the existing SARS-CoV-2 vaccines have substantially reduced the risk of severe disease and death up to this point. The induction of neutralizing antibodies (nAbs) by these vaccines leads to protection against both infection and severe disease. However, pharmacokinetic (PK) waning and rapid viral evolution degrade neutralizing antibody binding titers, leading to a rapid loss of vaccinal protection against infection occurring on the order of months after vaccination. Additionally, inter-individual heterogeneity in the strength and durability of the vaccine-induced neutralizing response to SARS-CoV-2 can create a further public-health risk by placing a subset of the population at risk. Here we incorporate the heterogeneity in inter-individual response into a pharmacokinetic/ pharmacodynamic (PK/PD) model to project the degree of heterogeneity in immune protection. We extend our model-based approach to examine the impact of evolutionary immune evasion on vaccinal protection. Our findings suggest that viral evolution can be expected to impact the effectiveness of vaccinal protection against severe disease, particularly for individuals with a shorter duration of immune response. One possible solution to immune heterogeneity may be more frequent boosting for individuals with a weaker immune response. We demonstrate a model-based approach to targeted boosting that involves the use of the ECLIA RBD assay to identify individuals whose immune response is insufficient for protection against severe disease. Our work suggests that vaccinal protection against severe disease is not assured and provides a path forward to reducing the risk to immunologically vulnerable individuals.

## Introduction

The rapid development of SARS-CoV-2 vaccines was an unprecedented achievement of modern science. Early reports suggested a high degree of vaccinal efficacy in preventing symptomatic disease [1–3], implying that the vaccines were effective at limiting transmission. This high level of vaccinal efficacy against infection raised the hope that the vaccines could be used to achieve herd immunity. However, this hope was soon undermined by waning antibody titers [4–7] and viral immune evasion [8–11], which predictably [12,13] led to rapid declines in vaccinal efficacy against infection [14,15].

At this point, a substantial body of evidence points to neutralizing antibody titers as a correlate of immune protection [16–18]. In a definitive meta-analysis, neutralizing antibody titers normalized to the mean convalescent titer (from the same study) demonstrated a strong nonlinear relationship that was predictive of reported vaccinal protection across a range of different vaccines [19]. The authors found a neutralizing antibody dose-response relationship between nAb titers and protection against infection, and a second dose-response relationship linking nAb titers to protection against severe COVID-19 outcomes. This relationship has held up across a range of studies [20,21], retaining strong predictive power even in the face of newly emerging variants [22–26]. Concomitant with waning neutralizing antibody titers and viral immune evasion, a number of studies have demonstrated a loss of vaccinal efficacy against severe disease (VE_s_) [27–30], contradicting a commonly held perception [31,32] that the observed durability of T-cell responses [33–35] would lead to prolonged vaccinal protection against severe disease.

In addition to population-level waning in nAb titers, there is significant inter-individual variation in the strength and durability of the nAb response [36–38]. In previous work, we have quantified this inter-individual heterogeneity by applying a mixed-effects modeling approach to published data characterizing SARS-CoV-2 nAb titers after time following infection [39]. Our results found a wide range of half-lives, with a 95% population interval ranging from 33-320 days. This wide range has significant implications for public-health strategy, as the existence of a subset of individuals who potentially lose immunological protection within a short span of time after infection also raises questions about the breadth and durability of vaccinal protection.

To explore this question in more depth, we use a population PK modeling approach to quantify the population heterogeneity in the durability of the nAb response as a result of vaccination. We then coupled this with the PK/PD dose-response relationships linking nAb titers to protection from mild and severe disease, in order to project the population-level heterogeneity. We examine population heterogeneity in vaccinal protection over time and in response to viral immune evasion. We further formulate and then evaluate a potential strategy for limiting risk for the vulnerable population based on the use of a personalized vaccine booster strategy.

## Results

### Population pharmacokinetic modeling of SARS-CoV-2 vaccinal immunity

We fitted a two-stage population mixed-effects model to nAb expansion and decay after vaccination. The selected model provides a good fit to nAb kinetics after the second vaccine dose based on the consistency of the observed data with the prediction intervals obtained from the model (Figure 1A). The model provides adequate estimation of population median and variation in all parameters (Table 1). We determined that there is a moderate correlation between T_in_ and k_p_. No significant correlations are observed between the initial nAb titer upon administration of the second vaccine dose and any of the model parameters. Additionally, there is no significant correlation between model parameters and age group either (Figure S1).

**Table 1.** Parameter values for fitted nAb kinetics model with standard errors (SE) and relative standard error (RSE).

| Parameter | Value | Units | Standard error | Relative standard error (%) |
| --- | --- | --- | --- | --- |
| *Fixed effects (median)* |  |  |  |  |
| $k_{p, \text{pop}}$ | 44.98 | IC50/days | 5.74 | 12.8 |
| $k_{el, \text{pop}}$ | 0.011 | 1/days | 0.0011 | 10.3 |
| $T_{in, \text{pop}}$ | 8.88 | days | 1.24 | 14.0 |
| *Standard deviation of the random effects* |  |  |  |  |
| $\omega_k$ | 0.61 | IC50/days | 0.093 | 15.3 |
| $\omega_{kel}$ | 0.47 | 1/days | 0.082 | 17.4 |
| $\omega_{T_{in}}$ | 0.66 | days | 0.12 | 18.8 |
| *Correlations* |  |  |  |  |
| $\text{corr}_{k, T_{in}}$ | -0.6 | | 0.15 | 24.2 |
| *Error model parameters* |  |  |  |  |
| b | 0.17 |  | 0.014 | 7.85 |

where $\text{corr}_{k, T_{in}}$ is the correlation between $k_p$ and $T_{in}$ , and b is the coefficient of proportional error.

**Figure 1:**
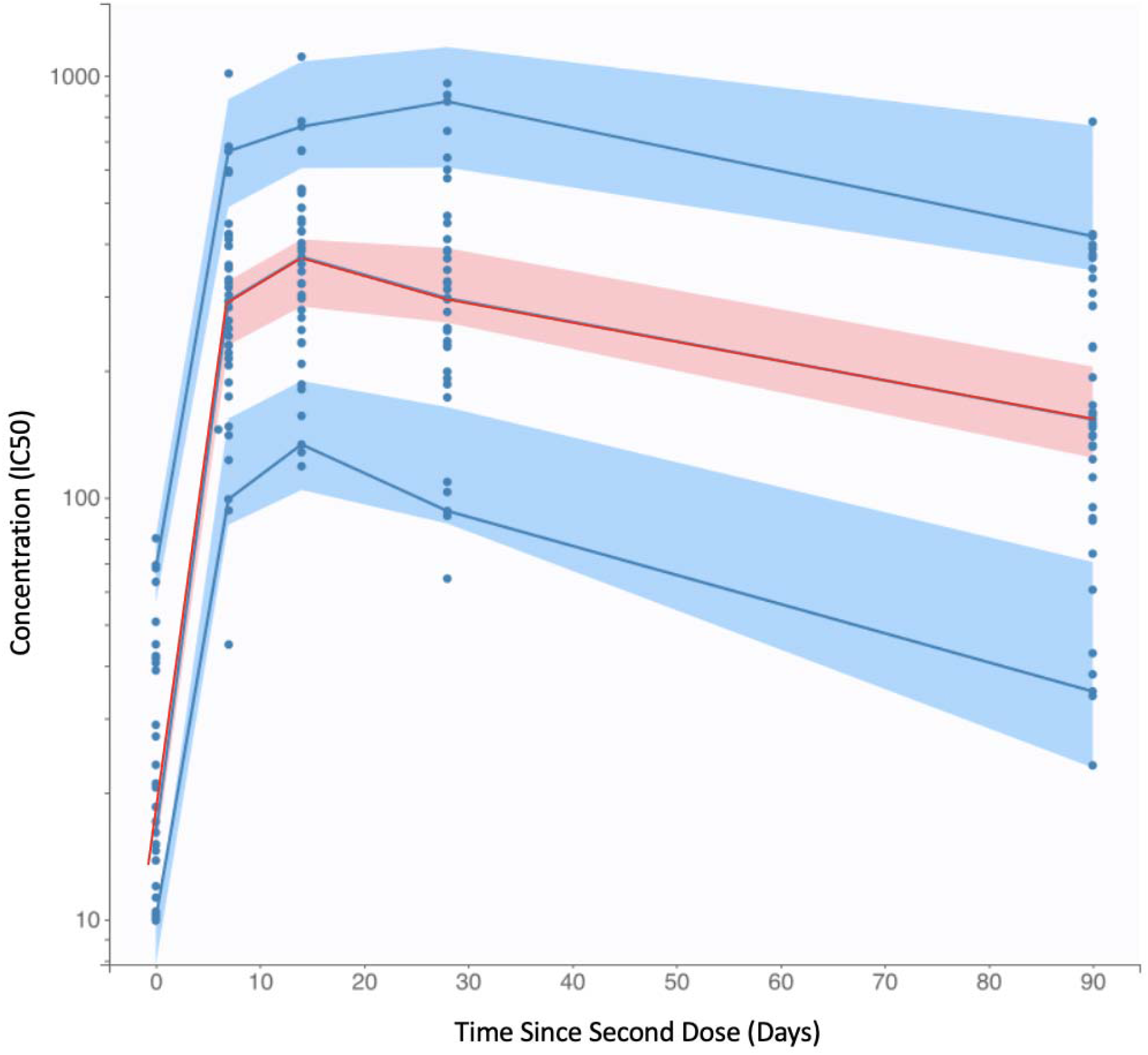
Visual predictive check (VPC) of nAb kinetics model fit with the 90% prediction intervals. The blue dots represent the observed clinical nAb kinetics data [4]. The red line represents the empirical 50^th^ percentile and the blue lines represent the empirical 10^th^ and 90^th^ percentiles. The shaded regions represent the model’s 90% prediction intervals for the 50^th^ percentile (pink) and 10^th^ and 90^th^ percentiles (blue). The empirical percentiles fall within the model’s prediction intervals, indicating good model agreement with the data.

The model was further validated by verifying agreement between the fitted parameter distributions and individual estimates and performing normality checks for random effects and residuals (Figures S2 - S4). We also note that the median half-life of nAbs significantly exceeds the median half-life of IgG antibodies (64 days vs 45 days, p = 0.005, Figure S5), which is consistent with affinity maturation occurring following vaccination.

### Peak nAb titer and half-life are heterogeneous in the general population

The population PK model reveals broad population heterogeneity in peak nAb titer and nAb half-life within the general (non-immunocompromised) population. The mean half-life for nAbs is 75 days (Figure 2A). For the mean, this translates to 29-fold waning of nAb titer one year after vaccination. In the upper 90^th^ percentile, the half-life is 127 days, implying 7.3-fold waning per year. However, the lower 10^th^ percentile has only a 36-day nAb half-life and experiences 1100-fold waning yearly. The mean peak nAb titer after vaccination is 469 (IC_50_), which is 4.4-fold of the mean convalescent plasma titer after infection (Figure 2B). The 90^th^ percentile for peak titer is 787 (IC_50_) (7.4-fold convalescent titer), and the 10^th^ percentile is 193 (IC_50_) (1.8-fold convalescent titer).

**Figure 2:**
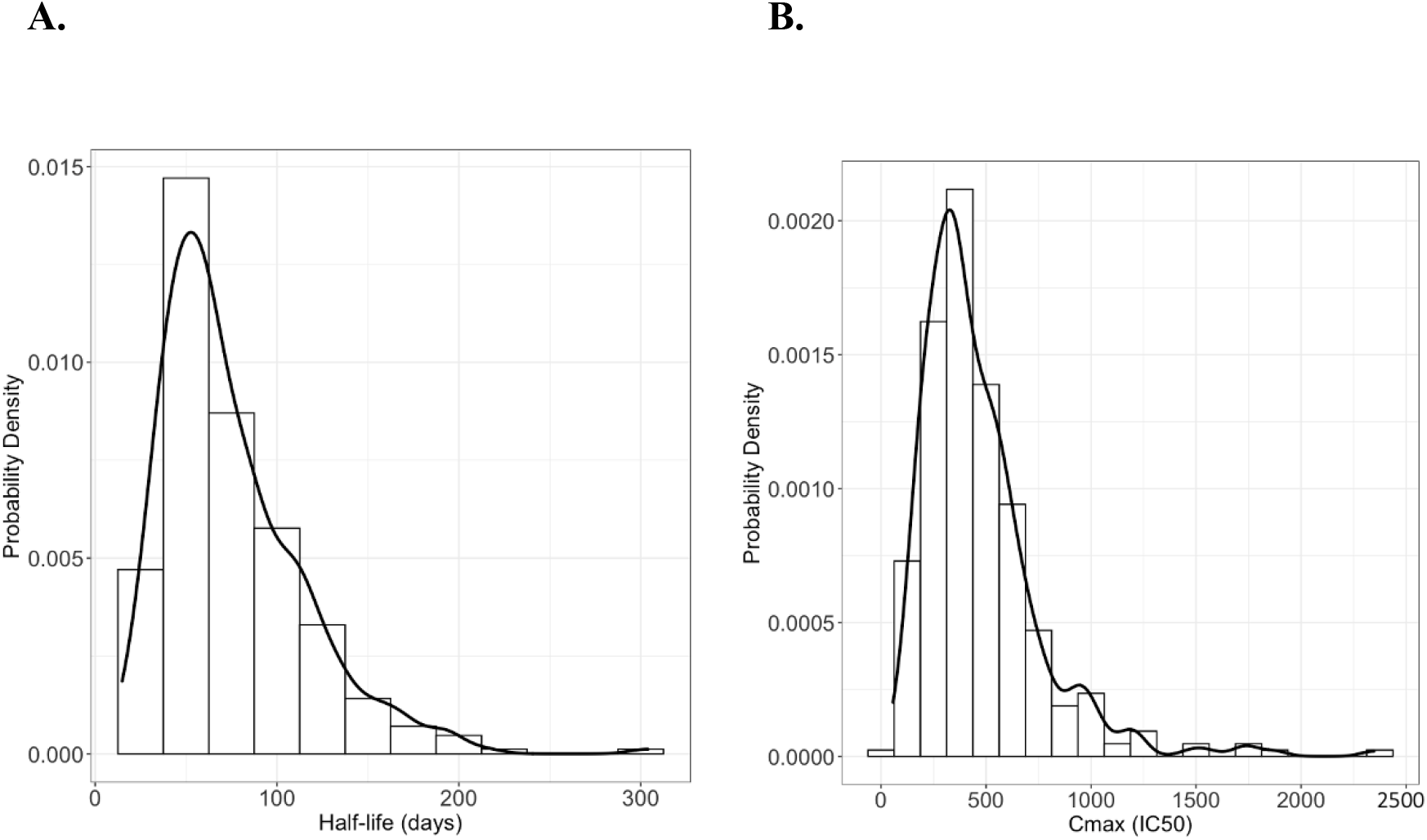
**A**. Distribution of model-fitted individual nAb half-lives in the study population after vaccination. **B**. Distribution of peak neutralizing titers after vaccination.

### Anti-SARS-CoV-2 nAbs wane after vaccination, with broad interindividual variability

In a model-simulated population, the heterogeneity in individual nAb PK parameters results in differences in nAb titers and persistence over time since infection (Figure 3). For individuals in the 50^th^ percentile, nAb titers are maintained above the peak convalescent level for about 4 months after vaccination. For the 10^th^ percentile, peak vaccine titers exceed the peak convalescent titer for 2 months, while for the 90^th^ percentile, vaccine titers remain above this threshold for approximately 10 months.

**Figure 3:**
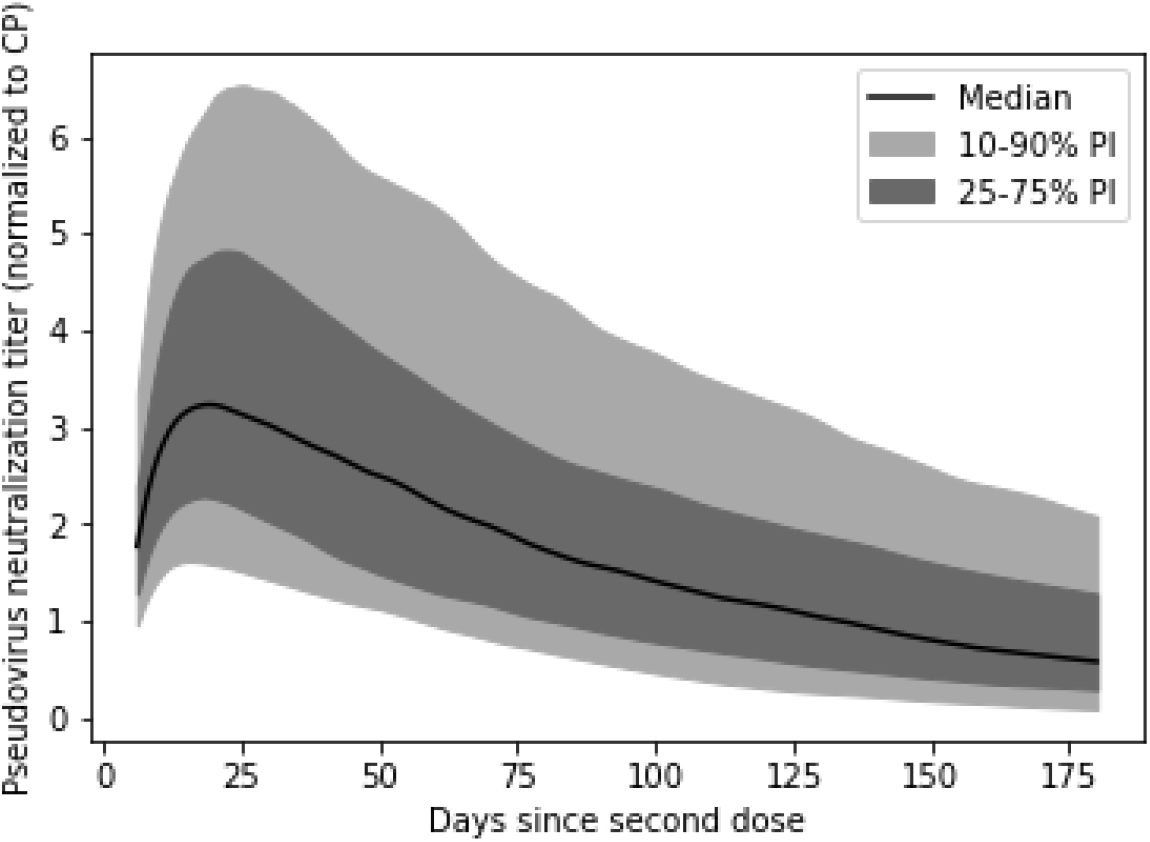
Pseudovirus neutralization titer over time by percentile. Titer is normalized to the mean peak convalescent level after SARS-CoV-2 infection.

### Variability in antibody persistence translates to functional differences in protection

Based on the simulated nAb kinetics, we estimated vaccine protection over time against COVID-19. As shown in Figure 4, vaccine protection over time varies based on differences in nAb PK. Protection from wild-type (WT) symptomatic (mild) disease (VE_m_) ranges from near-complete at the 90^th^ percentile to 90% in the 10^th^ percentile immediately after vaccination. As time progresses after vaccination, the variation increases – six months after the second dose, the 10^th^ percentile receives only 30% WT VE_m_, while the 90^th^ percentile retains 90% protection. Over this time interval, the population mean WT VE_m_ wanes from 94% to 67%. Across the board, protection from WT severe disease (VE_s_) is higher and more persistent than VE_m_ (Figure 4A). VE_s_ is near-complete across the population immediately after vaccination, and wanes to 75% after 6-months in the 10^th^ percentile. However, the mean VE_s_ for the population against WT remains 90% at 6 months. Thus, clinically significant differences in long-term vaccine efficacy are expected in the immunocompetent population, with potentially deadly consequences for those with poor nAb persistence.

**Figure 4:**
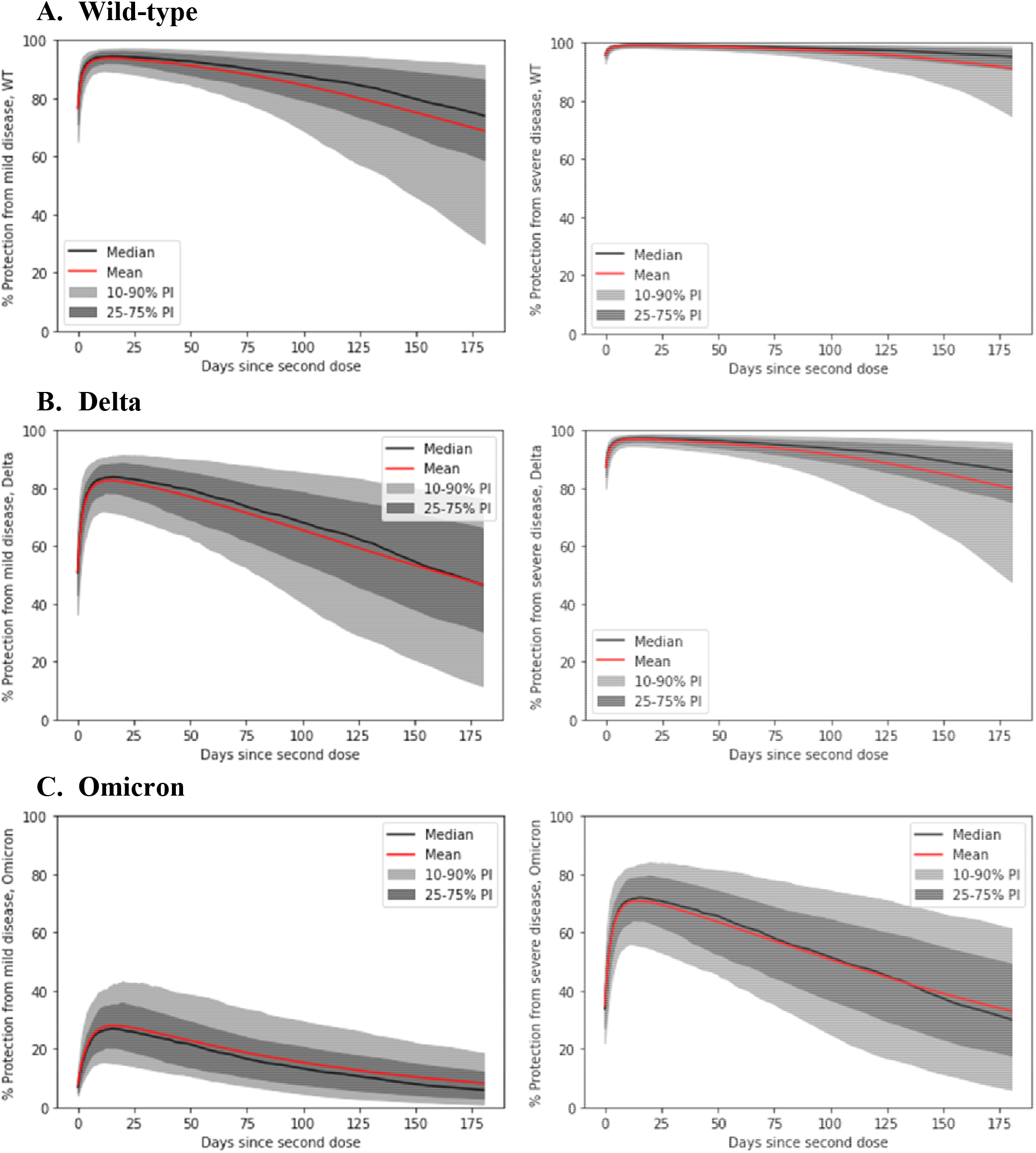
Protection from mild (left) and severe (right) COVID-19 by percentile over time since the second dose of the two-dose Moderna vaccination primary series. Protection is assessed for **A**. Wild type, **B**. Delta, **C**. Omicron.

### Immune evasion reduces vaccinal protection from severe disease

Although protection from severe disease remains relatively high throughout the population in the first 6 months after vaccination, immune-evading variants erode this protection, posing the risk of a rapidly-changing vaccinal immunity landscape as viral evolution continues. In Figure 4B, we demonstrate the challenge for vaccines when the more transmissible and modestly immune-evading delta variant emerged [40]. The mean VE_m_ disease immediately after vaccination dropped from about 94% to just over 80%. Although mean protection from delta severe disease is greater than 95% on average shortly after vaccination, it wanes more quickly than WT protection, reaching 80% after 6 months. Individuals with weaker vaccinal immunity are most impacted, with the 10^th^ percentile experiencing less than 50% delta VE_m_ 6 months after vaccination.

The strongly immune-evading omicron variant has had an even greater impact on vaccine efficacy (Figure 4C). On average, recently vaccinated individuals are conferred less than 30% protection from symptomatic omicron infections and 70% protection from severe disease. For the 10^th^ percentile, however, protection from omicron is minimal even shortly after vaccination – less than 20% against mild disease and approximately 55% against severe disease. Six months after vaccination, protection is poor across most of the population: virtually no one is expected to retain more than 30% protection from symptomatic omicron, while the mean and median levels of protection from severe omicron dip below 40%.

### Immune evasion erodes vaccine protection across the population

In Figure 5, we explore the relationship between degree of immune evasion – the fold-loss of nAb titer against an immune evading variant compared to the WT virus – and vaccine efficacy. Although mild disease is most impacted by immune evasion, severe disease protection is also predicted to be eroded, especially for strongly immune-evading variants. For example, a 7.5-fold loss of titer is expected to drop VE_m_ to 50% at three months post-vaccination, while a 50.8-fold loss of titer would reduce the median VE_s_ to 50% at three months.

**Figure 5.**
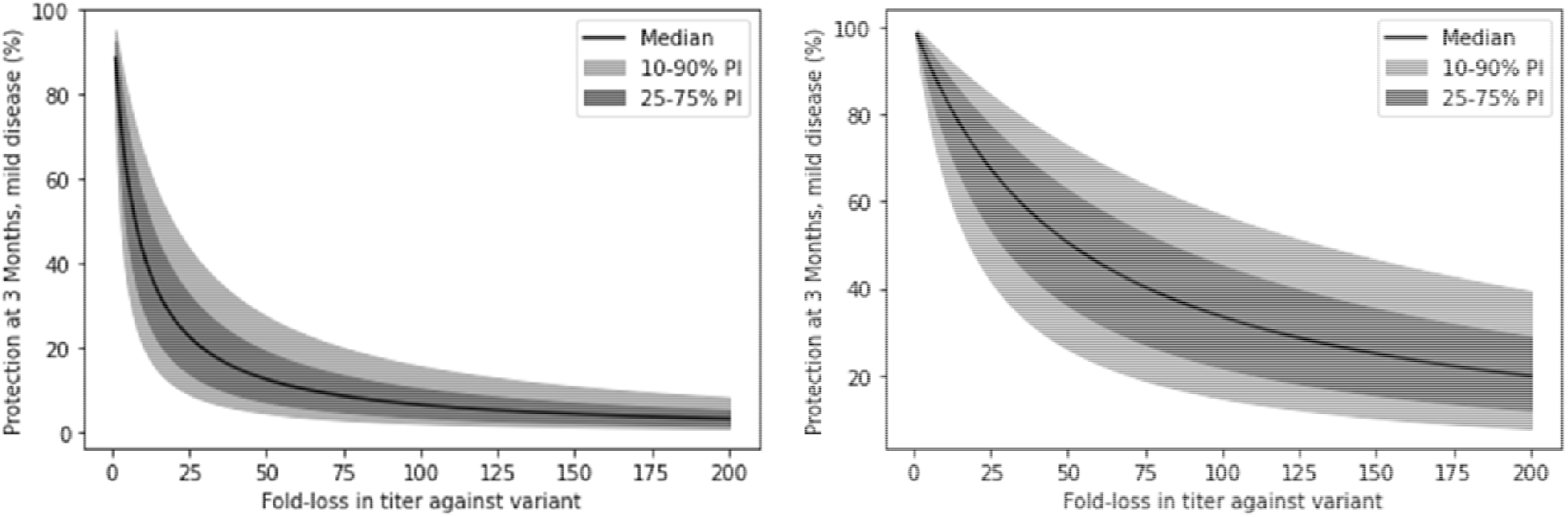
Protection from mild and severe disease at 3-months post vaccination as a function of immune evasion. Immune evasion is expressed as the fold-loss of vaccine serum titer against an immune-evading variant compared to WT pseudovirus.

### Model-based optimization of targeted booster doses

This analysis suggests that nAb waning and potency loss due to immune evasion contribute to substantial losses of protection over time, both from mild and severe disease. This waning is compounded by poorly persistent vaccinal immunity in a significant proportion of the immunocompetent population. Frequent revaccination resulting in boosting of titers is a potential solution to waning immunity and potentially immune evasion. Our results suggest that the optimal revaccination frequency may vary among individuals based on nAb PK. As nAb titer must be assessed by a live viral or pseudoviral assay, it is challenging to evaluate in a healthcare setting. In Figure 6, we demonstrate that RBD-binding IgG titer is a strong predictor of nAb titer above the level required for 90% protection from severe disease. For example, an optimized RBD-binding IgG titer can predict 90% protection from severe disease with 93% sensitivity and 72% specificity (Figure 6A). Thus, this more readily assessed metric could be used to identify patients with poor vaccinal protection who are good candidates for early revaccination.

**Figure 6:**
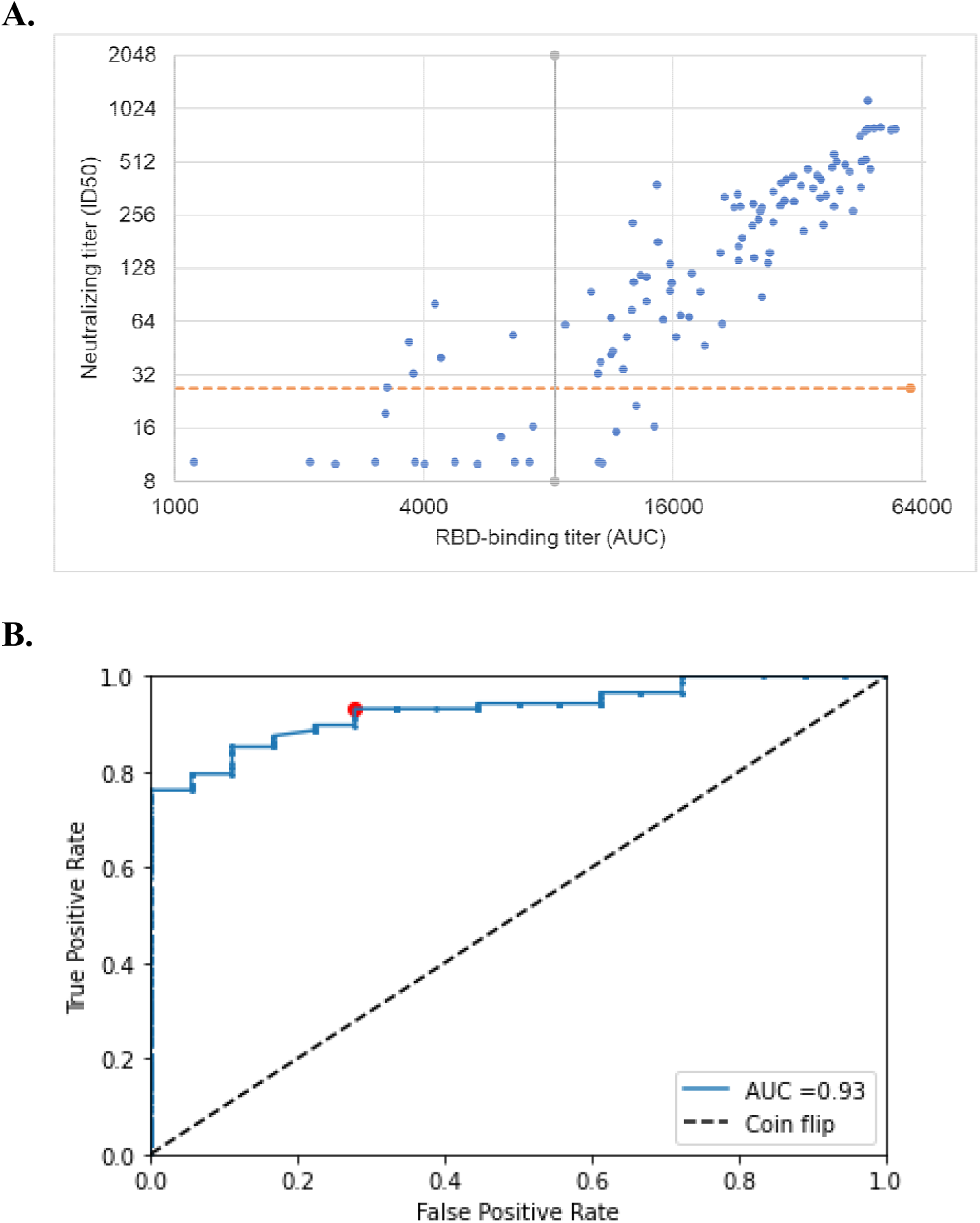
**A**. Scatter plot demonstrating correlation between RBD-binding titer and neutralizing titer. The red horizontal line represents the threshold for 90% protection from WT severe disease; vertical line represents a chosen threshold for targeted revaccination screening based on RBD-binding titer. **B**. ROC analysis on the dataset in panel A demonstrates that ECLIA RBD binding can predict whether vaccine protection is sufficient to provide 90% protection from severe disease. Red point represents the chosen threshold with 93% sensitivity and 72% specificity.

## Discussion

In this work, we have used a population PK/PD modeling approach to interrogate the impact of inter-individual heterogeneity on the degree and duration of vaccinal protection against both mild and severe disease. Our population model fit provided a good description of the data, as assessed by quality control metrics. The model showed broad heterogeneity in the degree and durability of nAb protection, with the 90% population interval (90% pi) for nAb half-life spanning 30 – 153 days, while the peak nAb level 90% pi spans 169 – 989 ID50.

The dataset used for this analysis was taken from a Moderna Phase 1 trial enrolling 34 participants, with immunocompromised status being an exclusion criterion for the trial. Thus, the broad heterogeneity observed is reflective of the diversity of outcomes that may be expected in the general population upon vaccination. Of particular concern, even though immunocompromised patients were specifically excluded from the underlying study, the outcomes for the 25^th^ percentile of the population and below are poor.. For example, omicron BA.1 VE_m_ and VE_s_ are both estimated to be less than 10% in this population at the six-month mark.

There are several limitations to our work, namely the small size of the Moderna study may not reflect all segments of the population and limits the power of the covariate analyses. The study excludes immunocompromised individuals, and thus our results reflect only the immunocompetent population. The Moderna study only covers the first three months after vaccination, which limits our ability to predict long-term nAb kinetics such as the possibility of biphasic decay. For this reason, we restricted our predictions to the first six months after vaccination and focused on variability in nAb kinetics during this timeframe.

Additionally, the results of the ROC analysis likely depend on a match between the circulating variant and the variant used to assess neutralizing titer and RBD-binding titer. Previous studies have shown that immune evading variants reduce the neutralizing potency of post-vaccination sera and that the relationship between binding and neutralizing titer varies [41]. Thus, as new variants emerge and sweep to dominance, this analysis will require repetition with pseudoviruses matched to the novel variants.

In the early days of the pandemic, there was much optimism expressed about the potential of vaccines to permit a return to normalcy, both in the popular press [42,43] and among public health authorities [44–46]. Much of this optimism was based on the persistence of the T-cell response in vaccinated individuals [35,47,48]. While T-cells remain durable even in the face of the newer immune-evading variants [35,49,50], this durability has not translated into lasting protection against infection or severe disease. At a mechanistic level, it is now known that T-cells are in fact infected by SARS-CoV-2 [51], and they undergo frank apoptosis during viral infection [51–53].

To the extent that neutralizing antibodies are the primary correlate of immune protection against SARS-CoV-2, our work makes several crucial points for public-health strategy. First, repeat annual dosing (at a minimum) of a SARS-CoV-2 vaccine may be required to provide population-level protection against severe disease. At present, the consequences of such a boosting strategy have not been fully explored in clinical trials-our work suggests that this is an urgent unmet medical need, and failure to keep providing boosters may lead to the loss of vaccinal protection against severe disease in the population. Understanding the impact of repeated boosting on nAb production as well as vaccinal side-effect profiles is crucial for enabling better use of the existing vaccines, which at present represent our only option for disease control.

Our work also points out a second unmet medical need, as many revaccination strategies may leave a significant portion of the immunocompetent population unprotected against severe disease. In the face of logistical constraints in vaccine production, a rational strategy would require a method to efficiently identify the subpopulation most in need of additional doses of vaccines. Our work further provides a basis for this prioritization. We have demonstrated an approach (based on previously published data [54]) that can be used to convert the ECLIA RBD-binding assay (which is commercially available and in broad use) into a personalized biomarker to determine the SARS-CoV-2 vaccine boosting interval for individuals in high-risk populations. The optimal threshold from our decision analysis provided 93% sensitivity and 72% specificity for predicting 90% protection from severe disease. Such an analysis would best be repeated in a larger prospective study to optimize the threshold for relevant variants and other experimental systems, but our findings here provide a clear basis for designing such a trial.

We recognize that repeated boosting with mRNA vaccines may present tremendous logistical hurdles on a global basis. However, the first step in solving a problem is to acknowledge its existence. Our work delineates the reality of the current situation-the path that we have chosen for attempting to coexist with SARS-CoV-2 will require us to either keep boosting the population at regular and frequent intervals, or risk losing vaccinal protection altogether. While manufacturing, tolerability, and compliance constraints may make this hard to achieve with the current vaccines, next-generation vaccines should be designed with this target product profile in mind. For example, room temperature-stable, nasally administered vaccines based on low-cost technologies would make it easier for us to achieve the goal of widespread and repeated vaccinal coverage.

A recurrent failing of the public-health strategy over the past two years has been to start with overly optimistic assumptions about the course of the pandemic, and to be slow to react to deviations from those assumptions. In fact, it was easy to predict that rapid viral immune evasion would be a problem [12], and it was easy to predict that the current vaccines alone would not bring the pandemic to an end [55]. At this point, our work suggests that we need to move quickly to bolster the protection provided by vaccines-by exploring dose and schedule effects of existing vaccines thoroughly, as well as by deploying next-generation vaccines. A failure to anticipate and hedge against waning vaccinal efficacy against severe COVID-19 outcomes could have grave consequences.

## Methods

### Population mixed effects model fit for neutralization potency and IgG levels

We use mixed-effect modeling to determine the population variability in kinetics of binding IgG and neutralizing antibodies generated by anti-SARS-CoV-2 vaccination. The clinical data is derived from a phase 1 trial of Moderna mRNA-1273 vaccine in 34 healthy adult participants who received two injections of vaccine at 100 μg [4]. We fitted the model to data collected starting on the administration date of the second dose in the two-dose vaccination series (day 0). The model’s initial concentration is set as the concentration on day 0.

To examine the rise and decay of longitudinal immune responses, we applied a two-stage model structure to the neutralizing potency and IgG level dataset. This model contains two phases: antibody production and memory phases. Exponential decay occurs in the whole process, while antibody production only occurs in the production phase. Zero-order or first-order production terms and possible correlation models were selected based on the Akaike information criterion and good parameter estimation (low standard error). Based on these criteria, the zero-order production model was selected for neutralizing potency and 1st-order production model was selected for IgG (See Table S2).

Zero-order production model:

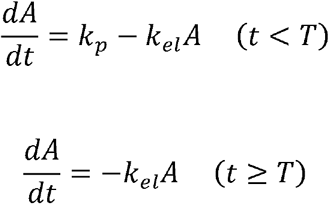

First-order production phase:

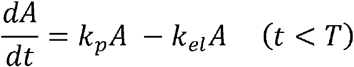

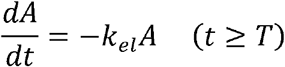

Where A is the antibody titer, k_p_ is the antibody production rate, k_el_ is the antibody elimination rate, and T is the duration of antibody production.

The population analysis was implemented in MonolixSuite. The residual error model is determined by whether PWRES (population weighted residuals), IWRES (individual weighted residuals) and NPDE (normalized prediction distribution errors) behave as independent standardized normal random variables. All parameters are lognormally distributed based on the default model structure. For lognormal distributed parameters, the predicted value (p) is represented by the following equation:

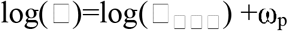

where □_□□□_ represents the fixed effect value, which is the median of the distribution in this case and ω the standard deviation of random effects, which is interpreted as the inter-individual variability. These specifications were evaluated using Monolix’s standard goodness of fit metrics including normality checks for distributions of random effects, scatter plots of population and individual weighted residuals, and distributions of individual values for model parameters (see Figures S2 - S4).

### Correlation and covariate analysis

Correlation and covariate analyses were implemented in Monolix to test the presence of correlations between model parameters and between model parameters and covariates. Age and initial conditions were assessed as potential covariates.

### Half-life and peak calculation

Both half-life and peak antibody titer for individuals in the two studies can be calculated directly from the structural model. Antibody half-life was calculated from individual decay rate as ln(2)/k_el_. The probability density functions of half-life were visualized for IgG and neutralizing potency from both datasets. Then, we compared the distribution of neutralizing potency and IgG to assess whether there is an affinity maturation. The peak antibody titer for individuals can be calculated by integrating the rate of increase of concentration 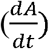 over the antibody production phase T_in_. The variability in individual half-life and peak titer values from the study population were visualized as distributions.

### Population variability in nAb titer over time

To visualize the population variability in nAb kinetics, we calculated the percentiles of nAb potencies over time in a bootstrapped synthetic population, considering both uncertainty of the population parameters and inter-individual variability in Simulx. Firstly, we formed a synthetic set of individuals by duplicating the study population 10 times. Parameters for these synthetic individuals were drawn from the uncertainty distributions computed by Monolix for each individual. Thus, the synthetic population reflects the uncertainty of the population parameters. The synthetic population was randomly drawn with resampling to generate a bootstrap population of 340 synthetic individuals. We used Simulx to simulate nAb titers over six months in this population and evaluated the percentile distribution of nAb titer at each day.

### Predicting vaccinal protection in the population based on nAb protection model

Based on the relationship between nAb titer and protection from mild or severe COVID-19 established by Davenport et al [19], we translated nAb kinetics in our simulated patient population to an expected level of protection over time. The simulated pseudovirus neutralization titers are normalized to COVID-19 convalescent plasma titers from a relevant dataset using the same assay and methodology [56]. We used the logistic titer-protection model to predict the level of protection based on our kinetic model’s simulated nAb titer normalized to convalescent plasma titer. This analysis reveals changes in risk of mild or severe COVID-19 over time by percentile in the population.

### Predicting vaccinal protection against SARS-CoV-2 variants

To estimate protection against SARS-CoV-2 delta and omicron variants, we assumed that these variants increase the nAb titer required for protection from mild and severe disease by a fixed multiple. The fixed multiple is the reduction in nAb potency against the variant compared to WT, as measured in a pseudovirus neutralization assay. According to the literature, delta reduced nAb potency after the Moderna two-dose vaccine series by 3.2-fold relative to WT, whereas omicron reduced potency 43-fold [40].

### Assessing IgG titer as a predictor of protective nAb titer

To determine whether IgG titer could be used to predict nAb titers above the EC_90_ for protection from severe WT disease (27 ID_50_), we performed a receiver operating characteristic (ROC) analysis. The ROC curve is formed by calculating the sensitivity and specificity of various IgG titer thresholds for predicting nAb titer above 27 ID_50_. The dataset for this analysis is sourced from Pegu et al [54], which was also produced by Moderna.

## Data Availability

All data produced in the present study are available upon reasonable request to the authors.

## Supplement

**Figure S1.**
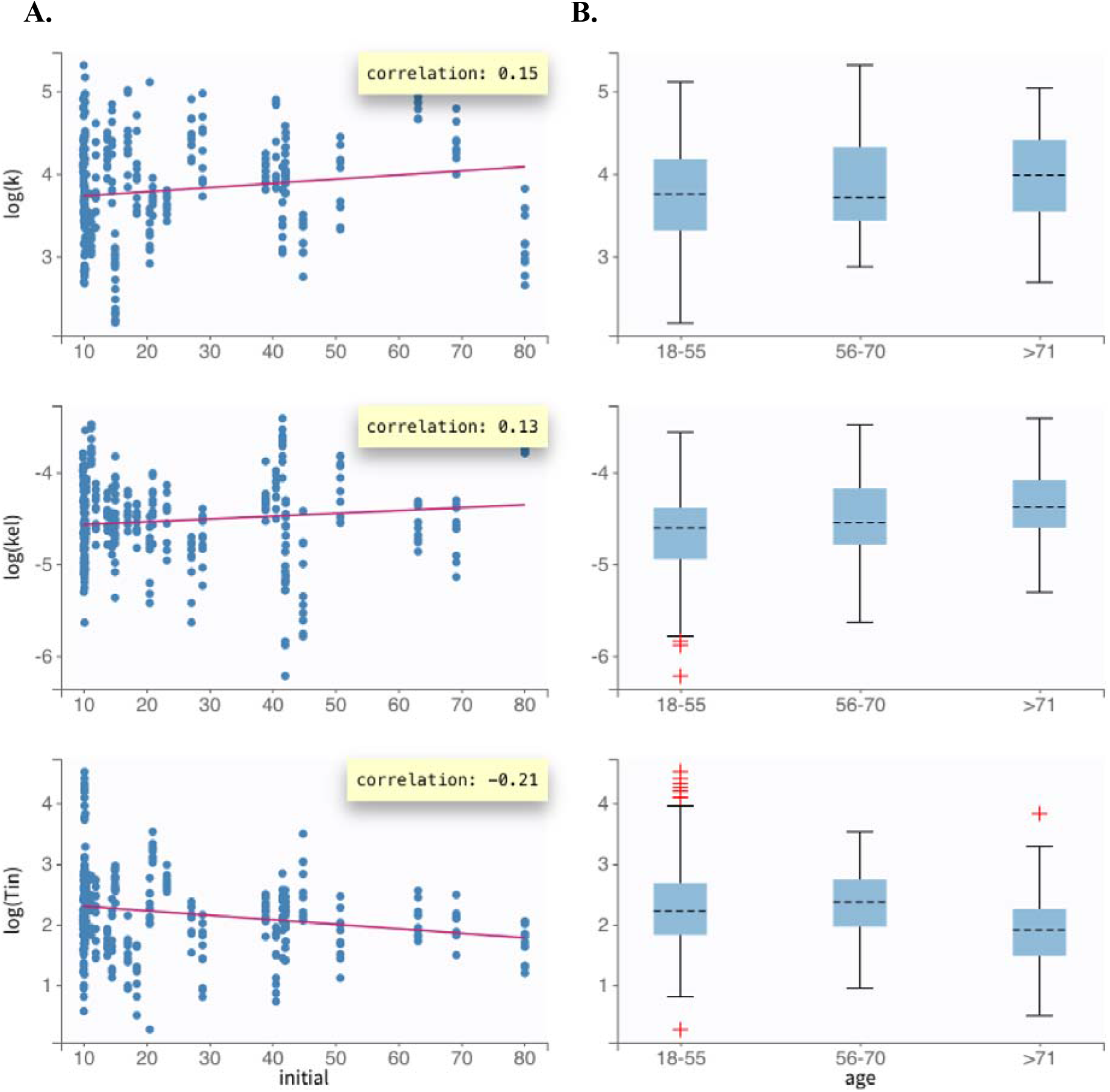
Diagnostic plot assessing correlations between individual parameters and possible covariates. **A**. The initial condition, the nAb titer at day 0, and **B**. age were assessed as potential covariates. No significant correlations were found.

**Figure S2:**
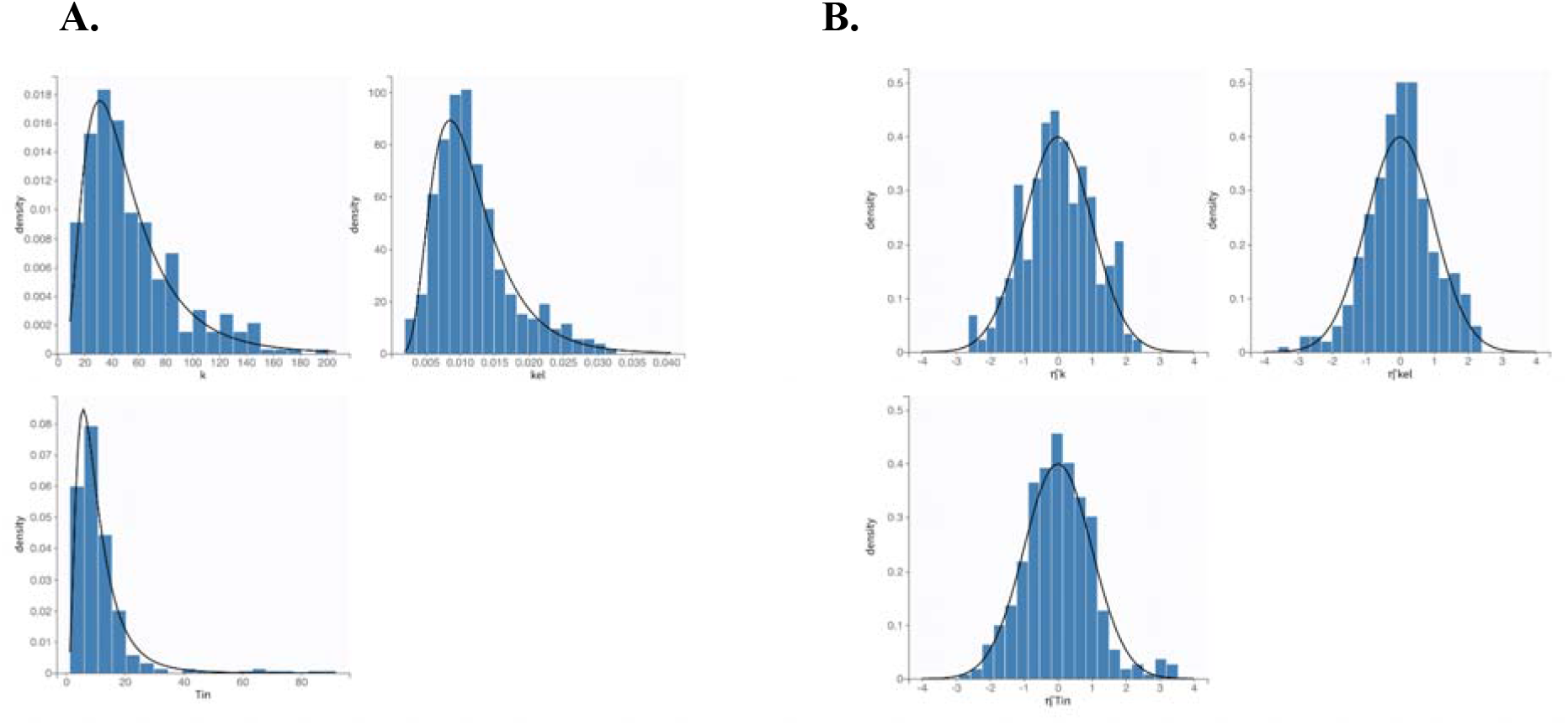
Probability distribution of individual parameter estimates and random effects for the nAb kinetics model. A) Probability distribution of individual parameters. The histogram plots represent the empirical distribution. The black line represents the theoretical distribution defined in the statistical model, which is a log-normal distribution for each parameter. B) Probability distribution of standardized random effects. The histogram plots represent the empirical distribution. The black line represents the theoretical distribution defined in the statistical model, which is a normal distribution for each random effect.

**Figure S3.**
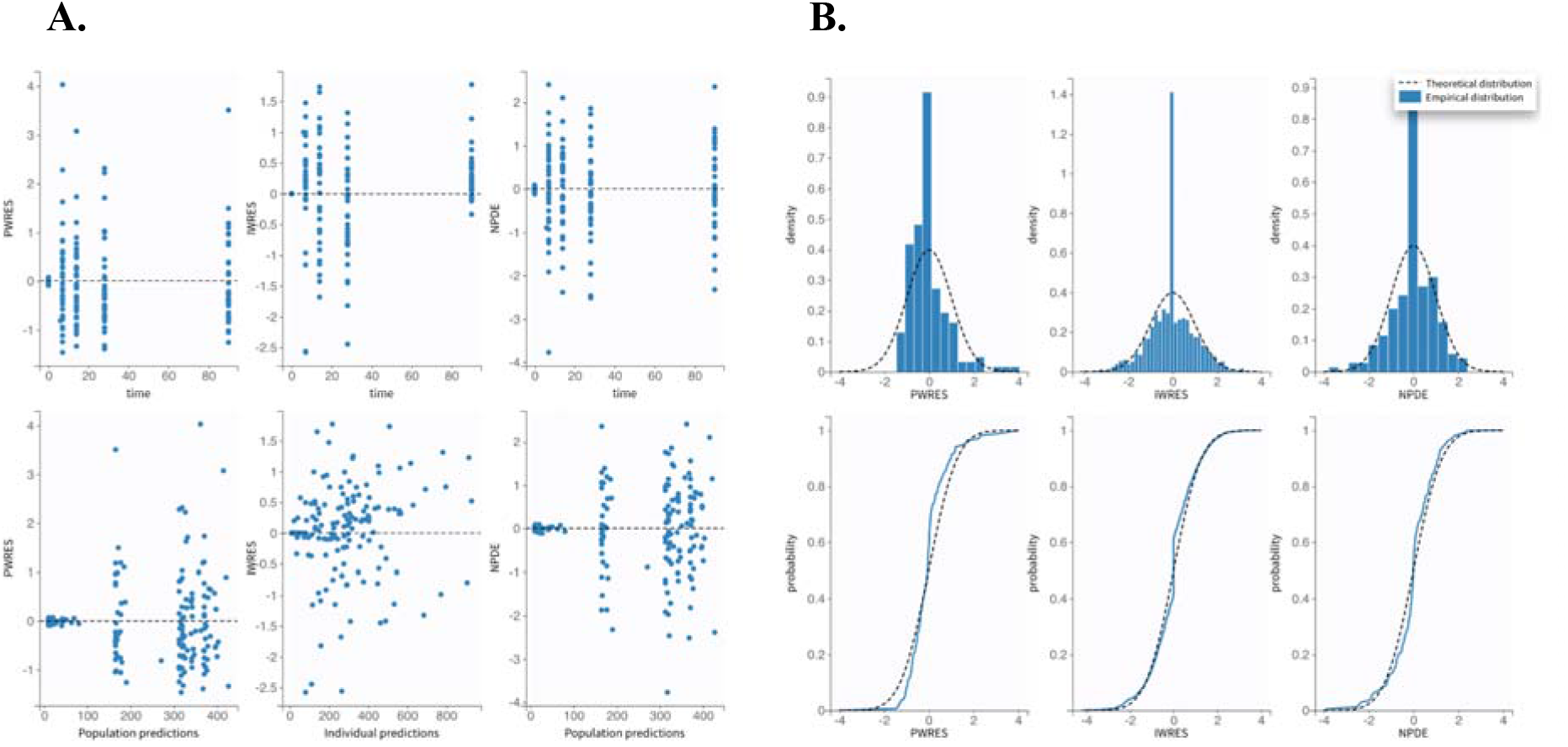
Goodness-of-fit analysis for nAb kinetics model residuals. **A**. Scatter plot of the residuals. These plots display the PWRES (population weighted residuals), the IWRES (individual weighted residuals), and the NPDEs (normalized prediction distribution errors) as scatter plots with respect to time and prediction. Residuals should be randomly scattered around the x-axis, which confirms suitability of the proportional error model. **B**. Distributions of the residuals. Empirical and theoretical probability density function (PDF) of the PWRES, IWRES and NPDE are shown in the top of the panel. Empirical and theoretical cumulative distribution function (CDF) are at the bottom. This normality check confirms suitability of the error model.

**Figure S4.**
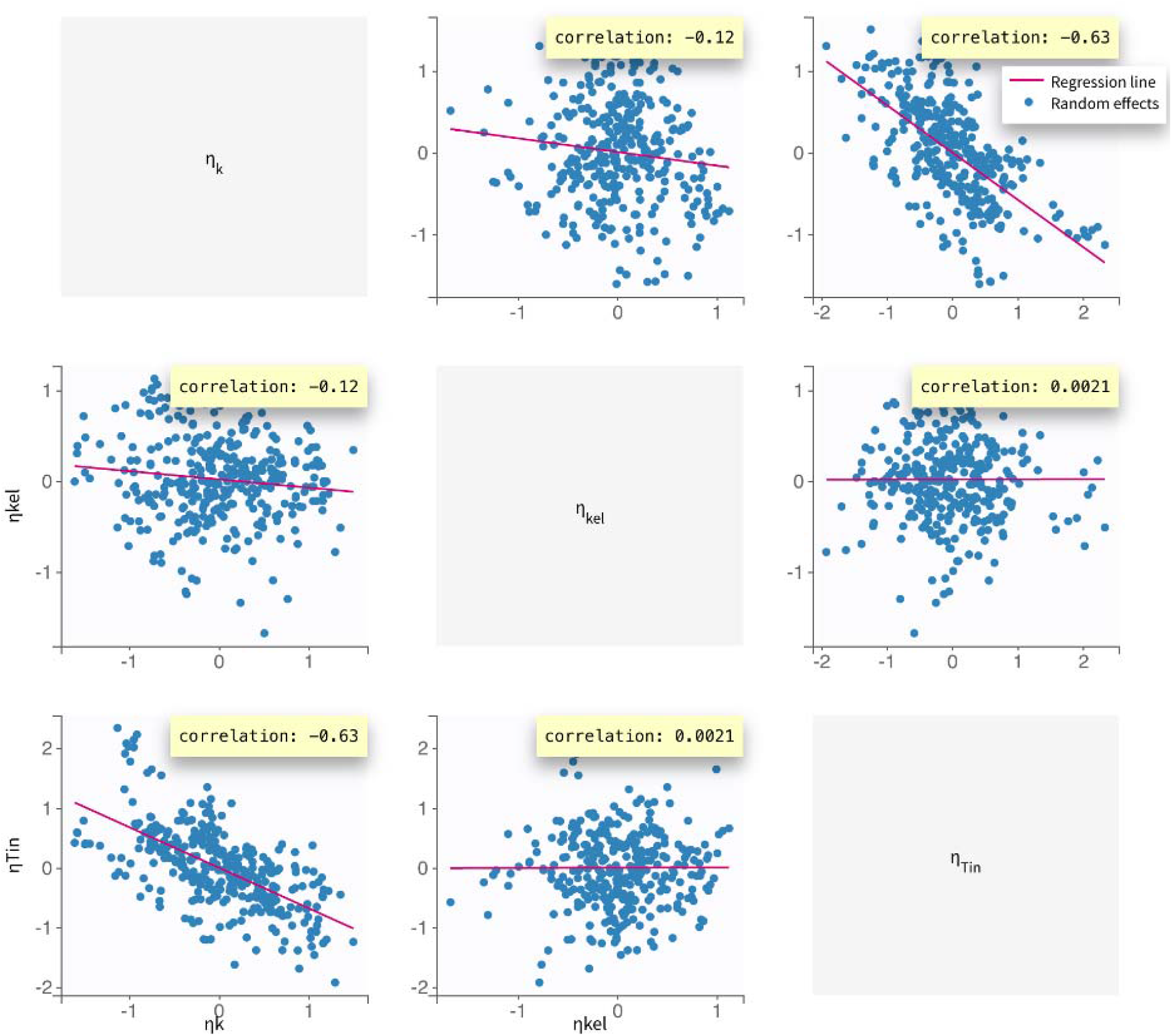
Correlation between parameters. There is a moderate correlation between Tin and k_p_.

**Figure S5:**
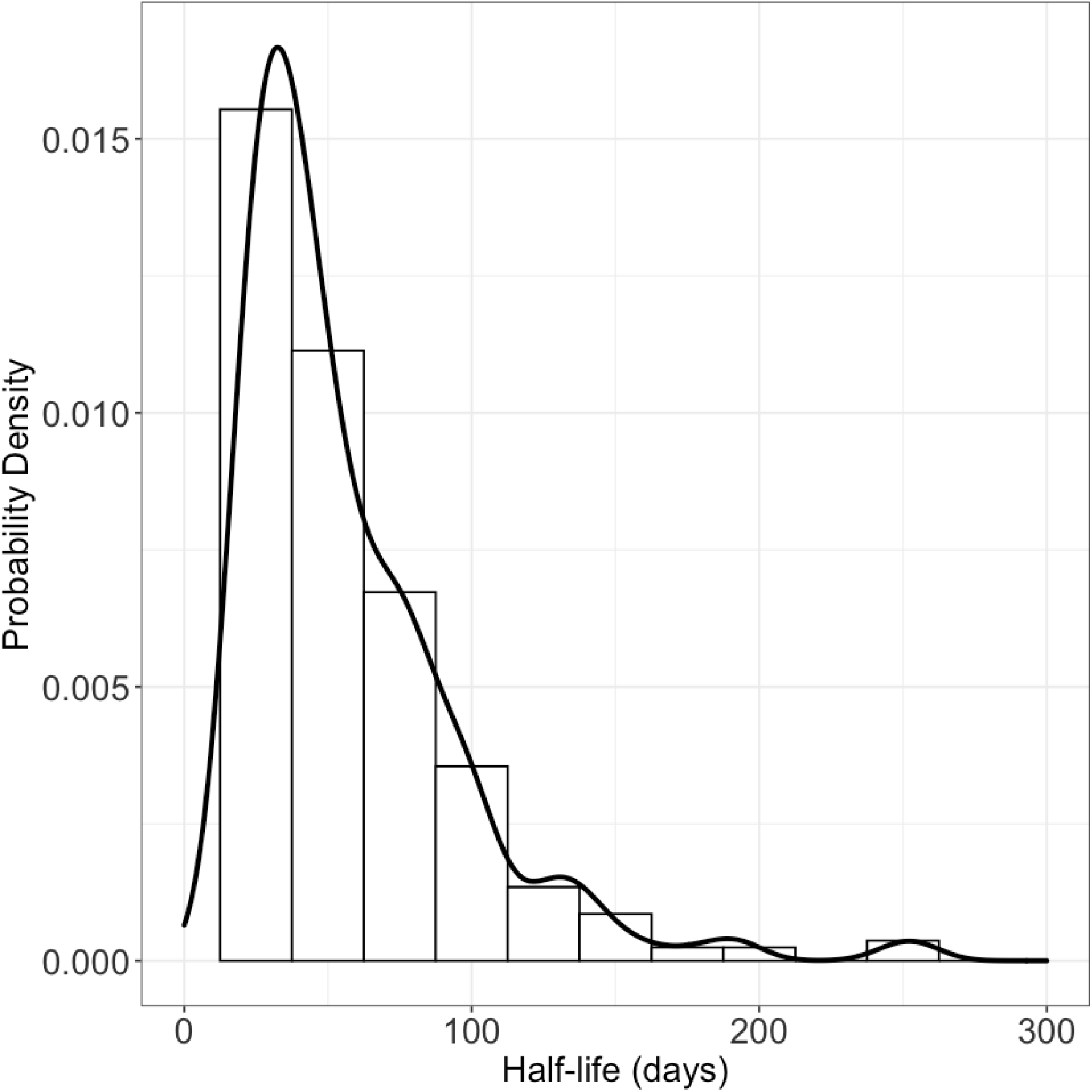
Probability distribution of half-lives for vaccine-induced IgG for individuals in the study population.

**Figure S6.**
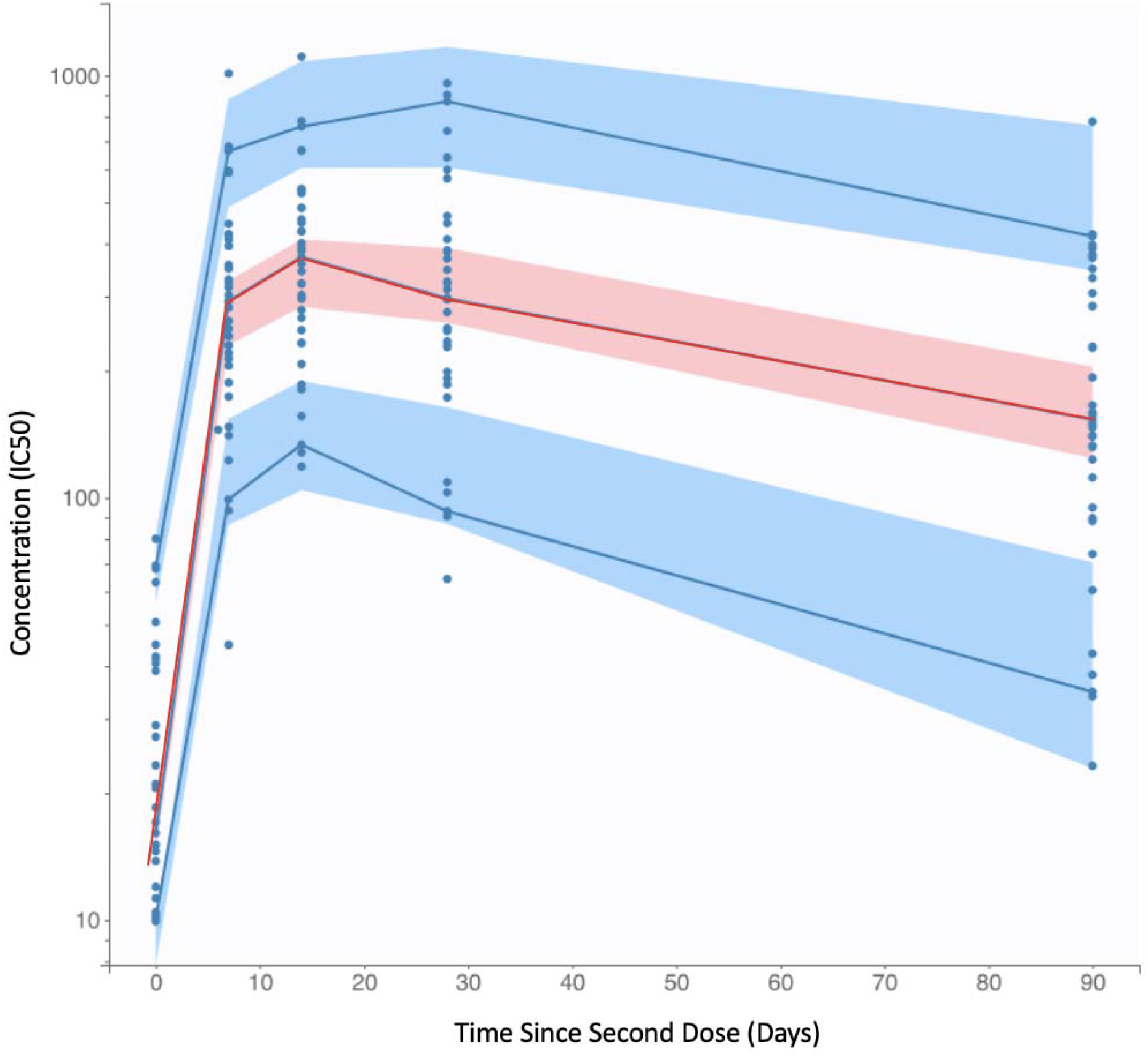
VPC of IgG kinetics model fit with 90% prediction intervals. Blue dots represent published data characterizing SARS-CoV-2 IgG titers following vaccination. The red line represents the empirical 50^th^ percentile and the blue lines represent the empirical 10^th^ and 90^th^ percentiles. The shaded regions represent the model’s 90% prediction intervals for the 50^th^ percentile (pink) and 10^th^ and 90^th^ percentiles (blue).

**Table S1.**
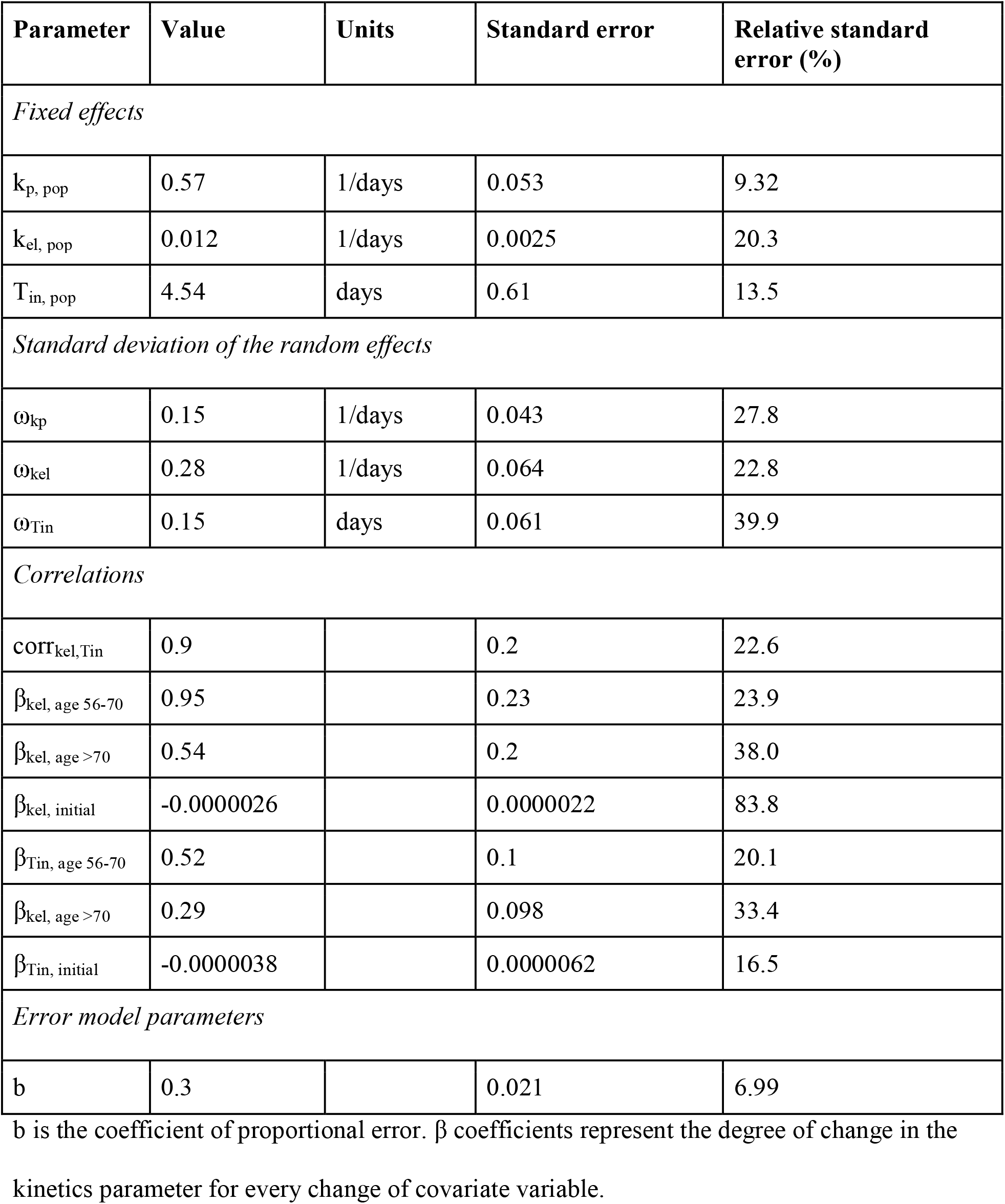
Parameter values for fitted IgG kinetics model with standard errors (SE) and relative standard error (RSE).

**Table S2.** AIC values for prospective model structures.

| **Antibody type** | **Model** | **AIC** |
| --- | --- | --- |
| nAbs | 0-order production | 1842.04 |
| nAbs | 1<sup>st</sup>-order production | 1894.46 |
| IgG | 0-order production | 4278.69 |
| IgG | 1<sup>st</sup>-order production | 4261.57 |

**Table S3.** Comparison of model-predicted vaccine efficacy over time with clinical data.

| **Variant** | **Time since second dose (days)** | **VEm, model estimate (%)** | **VEm, clinical (%**<br>**95% CI) [40]** |
| --- | --- | --- | --- |
| Delta | 14-90 | 76.4 | 82.8 (69.6, 90.3) |
| Delta | 90-180 | 57.2 | 63.6 (51.8, 72.5) |
| Omicron | 14-90 | 22.6 | 30.4 (5.0, 49.0) |
| Omicron | 90-180 | 11.9 | 15.2 (0.0, 30.7) |

